# COVID-19 in hemodialysis (HD) patients: Report from one HD center in Wuhan, China

**DOI:** 10.1101/2020.02.24.20027201

**Authors:** Yiqiong Ma, Bo Diao, Xifeng Lv, Wei Liang, Jili Zhu, Lei Liu, Sihao Zhang, Bo Shen, Huiming Wang

## Abstract

**Importance:** The outbreak of highly contagious COVID-19 has posed a serious threat to human health, especially for those with underlying diseases. However, Impacts of COVID-19 epidemic on HD center and HD patients have not been reported.

**Objective:** To summery an outbreak of COVID-19 epidemic in HD center.

**Design, Setting, and Participants:** We reviewed the epidemic course from the first laboratory-confirmed case of COVID-19 infection on January 14 to the control of the epidemic on March 12 in the HD center of Renmin Hospital of Wuhan University. Total 230 HD patients and 33 medical staff were included in this study

**Exposures:** COVID-19.

**Main Outcomes and Measures:** Epidemiological, clinical, laboratory, and radiological characteristics and outcomes data were collected and analyzed. 19 COVID-19 HD patients, 19 non-COVID-19 HD patients and 19 healthy volunteers were enrolled for further study about the effect of SARS-CoV-2 infection on host immune responses.

**Results:** 42 out of 230 HD patients (18.26%) and 4 out of 33 medical staffs (12.12%) were diagnosed with COVID-19 from the outbreak to March 12, 2020. 13 HD patients (5.65%), including 10 COVID-19 diagnosed, died during the epidemic. Only 2 deaths of the COVID-19 HD patients were associated with pneumonia/lung failure. Except 3 patients were admitted to ICU for severe condition (8.11%), including 2 dead, most COVID-19 diagnosed patients presented mild or none-respiratory symptoms. Multiple lymphocyte populations in HD patients were significantly decreased. HD patients with COVID-19 even displayed more remarkable reduction of serum inflammatory cytokines than other COVID-19 patients.

**Conclusions and Relevance:** HD patients are the highly susceptible population and HD centers are high risk area during the outbreak of COVID-19 epidemic. HD Patients with COVID-19 are mostly clinical mild and unlikely progress to severe pneumonia due to the impaired cellular immune function and incapability of mounting cytokines storm. More attention should be paid to prevent cardiovascular events, which may be the collateral impacts of COVID-19 epidemic on HD patients.

## Introduction

Since December 2019, the initially outbreak of 2019 novel coronavirus disease (COVID-19) in Wuhan city has spread rapidly worldwide, becoming a serious pandemic [1-6]. Epidemiological survey showed that COVID-19 patients with underlying conditions such as diabetes, hypertension, cardiovascular disease or the elderly are not only susceptible, but also often more serious [7-10]. However, as far as we know, the impact of COVID-19 epidemic on chronic kidney disease patients, especially HD patients has not yet been reported. Considering the large population size of HD patients (There are 7184 registered patients receiving HD treatment in 61 centers in Wuhan city.), high concentration of patients in HD center, and the compromised immune function of uremic patients [11], the situation of HD patients under the COVID-19 epidemic should not be ignored.

In this study, we reviewed an outbreak of COVID-19 in one HD center in Renmin Hospital of Wuhan University, one of the largest hospitals located in the downtown of Wuhan city. A cluster of HD patients contracted COVID-19 were surveyed and followed until the day on March 12, 2020. The epidemiological, clinical, laboratory, and radiological characteristics, and outcomes of some of these patients were reviewed. We expect our findings will shed light on the appropriate management of the HD center and HD patients in face of COVID-19 or other similar epidemic emerging.

## Methods

### Study design and participants

We reviewed the epidemic course from the first laboratory-confirmed case of COVID-19 infection on January 14 to March 12 in the HD center of Renmin Hospital of Wuhan University. Total 230 HD patients and 33 medical staff were included in this study. Diagnosis of COVID-19 pneumonia was based on the New Coronavirus Pneumonia Prevention and Control Program (5^th^ edition valid from Feb 4, 2020 to Feb 17, 2020, 6^th^ edition valid from Feb 18, 2020 to Mar 4, 2020, 7^th^ edition valid from Mar 4, 2020 until now) published by the National Health Commission of China [12-14]. In the 5^th^ edition, the suspect case of COVID-19 is defined as the one has the epidemiological history or clinical presentations of fever, respiratory symptoms, or decreased white blood cells or lymphocytes count. The clinical diagnosed case is recognized when the suspect case displays the imaging features of pneumonia. The confirmed case is identified if the suspected cases or clinical diagnosed case is positive in pathogenic test. In the 6^th^ and 7^th^ edition, the definition of clinical diagnosed case is removed. The suspect case should meet the criteria of epidemiological history and clinical presentations. In the presence of pathogen evidence, the suspect case should be confirmed diagnosis of COVID-19. We followed up all patients and collected the related clinical data. The study protocol was approved by the Ethics Committee of Renmin Hospital of Wuhan University (WDRY2020-K064). Written informed consent was waived by the Ethics Commission of the hospital for emerging infectious diseases.

### Data collection

The medical records of all participants were analyzed by the research team of the Department of Nephrology, Renmin Hospital of Wuhan University. Epidemiological, clinical, laboratory, and radiological characteristics and outcomes data were obtained with data collection forms from electronic medical records or specific data questionnaire. Some data were retrieved from the Hubei Province kidney disease quality control information platform. The data were reviewed by a trained team of physicians. Information collected including demographic data, medical history, underlying comorbidities, symptoms, signs, blood tests and chest computed tomographic (CT) scans. During the period of follow up, the odd episode of death were recorded and the presumed cause of death were carefully evaluated by the research team, based on the time, place and clinical manifestations.

### Virologic Studies

The SARS-CoV-2 detection was done by real-time PCR (RT-PCR) as described previously [15]. Briefly, nasopharyngeal swab samples of participants were collected for SARS-CoV-2 test with the detection kit (Bioperfectus, Taizhou, China). The ORF1ab gene (nCovORF1ab) and the N gene (nCoV-NP) were used for real-time RT-PCR according to the manufacturer’s instructions. Reaction mixture were prepared and RT-PCR assay was then performed under the following conditions: incubation at 50 °C for 15 minutes and 95 °C for 5 minutes, 40 cycles of denaturation at 94°C for 15 seconds, and extending and collecting fluorescence signal at 55 °C for 45 seconds.

### Cellular immune profiling and cytokines measurement

To explore the effect of SARS-CoV-2 infection on host immune responses, we recruited 19 COVID-19 HD patients and 19 non-COVID-19 HD patients for further blood sample collection with their permission. 19 Healthy volunteers were simultaneously enrolled. Peripheral blood mononuclear cells (PBMCs) and sera were isolated. PBMCs were stained with a BD Multitest IMK Kit (Cat340503, BD Biosciences) for analyzing the cell frequency of total T, CD4^+^ T, CD8^+^ T, B and NK cells in healthy controls and patients. The stained cells were acquired on a LSR Fortessa Cell Analyzer (BD Biosciences) and data analyzed using the FolwJo software (TreeStar). Serum level of a panel of cytokines covering IL-4, IL-6, IL-10, TNF-α, IFN-γ were assayed using Human Cytokine Standard 27-Plex Assays panel and the Bio-Plex 200 system (Bio-Rad, Hercules, CA, USA) for all patients according to the manufacturer’s instructions. All experimental procedures were completed under biosafety level II plus condition.

### Statistical Analysis

The measured data were using median and interquartile range (IQR) values and compared using independent group *t* test. Enumeration data were described as number (%). All statistical analyses were performed using SPSS (Statistical Package for Statistical analysis), and a *P* value of less than 0.05 is considered as significant difference.

## Results

230 patients and 33 staff in our HD center were included in this study. The dynamic course of COVID-19 epidemic from emerging to development is schematically presented in Figure 1A. The first COVID-19 patient was diagnosed on January 14, and the second diagnosed patient appeared on January 17. Since January 21, patients with COVID-19 had been quarantined and all medical staff had been asked to upgrade their personal prevention and protection, which including wearing full protective gear such as waterproof disposable gown, cap, gloves, face shield, and N95 face mask, and more rigorous cleaning and disinfection. On February 4, 2 new patients and 3 medical staffs were further confirmed with COVID-19. Therefore, the HD center decided to screen all patients and staffs on February 4 with chest CT and optional blood test. On February 10, there were 29 cases being diagnosed with COVID-19 including 29 HD patients and 1 medical staff. On February 13, 2020, 4 more COVID-19 cases were confirmed in HD patients. Since then, until the first screening was fully completed on Feb 17, 2020, no new COVID-19 case occurred. In order to find out the potential infected case in incubation period, we launched the second round of screening from Feb 22, 2020 to Mar 3, 2020, and the third round of screening from Mar 3, 2020 to Mar 12, 2020. There were 3 cases in 2^nd^ screening, and 2 cases in 3^rd^ screening, being confirmed diagnosis of COVID-19.

**Figure 1.**
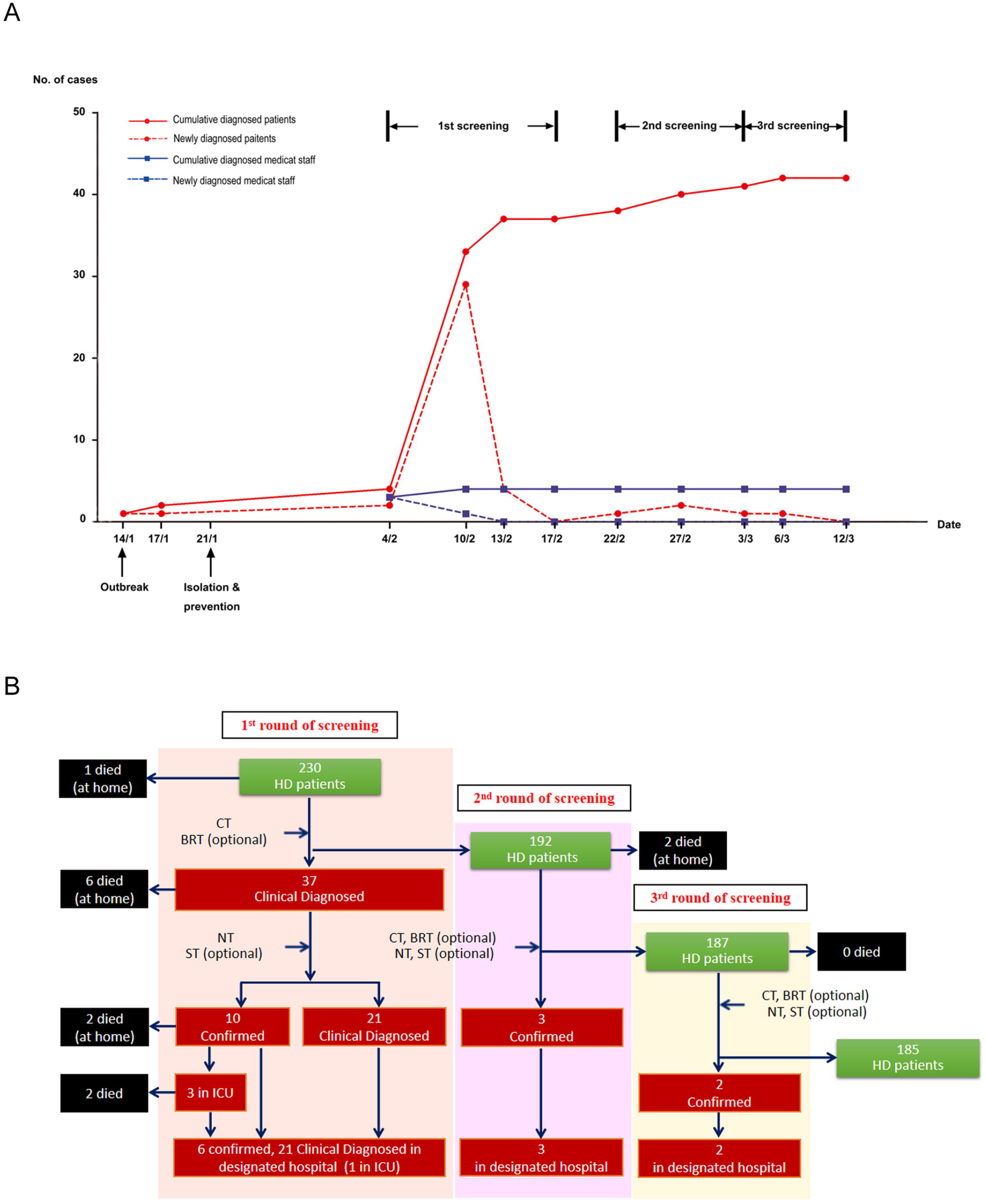
**A:** The schematic diagram of dynamic course about COVID-19 epidemic in our HD center. The first COVID-19 patient was diagnosed on January 14. The second patient was diagnosed on January 17. The personal prevention and protection of medical staff was upgraded on January 21. 2 new patients and 3 medical staff were confirmed with COVID-19 on February 4. 29 HD patients and 1 medical staff were diagnosed on February 10. 4 new HD patients were diagnosed with COVID-19 on February 13. 3 cases were confirmed in the 2^nd^ round of screening, and 2 cases were confirmed in the 3^rd^ round of screening. **B:** The management flow and the outcomes of the cluster in the epidemic. In the 1^st^ round of screening, 37 patients were diagnosed with COVID-19 in our center. Among them, 10 patients had died. 3 infected patients were admitted to ICU for severe conditions and 2 had died. In the 2^nd^ round of screening, 3 patients were confirmed. In the 3^rd^ round of screening, 2 patients were confirmed. All of the diagnosed patients were transferred to the designated hospitals.

During the period of screening, all infected patients and staffs were classified, quarantined, or transferred to designated hospital according to the government instruction. Figure 1B summarized the management flow and the outcomes of the cluster. It showed that totally 42 (18.26%) patients were diagnosed with COVID-19, 13 patients have died since the epidemic outbreak, including 10 patients contracted COVID-19. Among the infected patients, 3 were admitted to ICU for severe conditions and 2 had died later, the left one is in stable condition. We launched 3 rounds of screening to find out the infected patients in the HD center. The first round started on Feb 4, 2020 and closed on Feb 17, 2020, with 37 patients being clinical diagnosed of COVID-19. There were 10 patients were confirmed the diagnosis of COVID-19 based on the positive test of SARS-CoV-2 nucleic acid on Nasopharyngeal swab. All of these 31 patients were transferred to the designated hospitals, and the remaining stayed in our hospital under indicated quarantine or isolation, followed by the 2^nd^ screening, beginning on Feb 22, 2020. Three patients were confirmed diagnosis and other 2 patients had died in this phase. The 3^rd^ round of screening was carried out on 187 patients and found out 2 patients contracted COVID-19. As of Feb 14, 2020, there were 185 HD patients remaining in our hospital and 32 in the designated hospital. All of the deaths were followed and reviewed by our research team. Except 2 patients died in ICU, the other 11 cases died at home and showed no obvious symptoms of pneumonia, their inferred cause of death was heart failure, hyperkalemia and cerebrovascular disease, and none of them succumbed to severe pneumonia based on the clinical manifestations (Figure 1B).

Demographic data of 42 COVID-19 HD patients were summarized in Table 1. Among 15 confirmed diagnosed COVID-19 patients, 10 were male and 5 were female, with median age of 71 years. 27 clinical diagnosed COVID-19 patients contain 15 male and 12 female, with median age of 61 years. Hematology abnormalities such as lymphocytopenia and thrombocytopenia are common in COVID-19 patients as previous reports [7, 8, 16, 17]. These features were also present in the COVID-19 contracted HD patients (Table 2). With respect to the clinical symptoms in HD patients with COVID-19, we found that symptoms of fever, fatigue, dry cough, chest pain, as well as nausea, were either not common (Table 2). Radiological detection by chest CT scan showed that 2 (13%) confirmed diagnosed patients and 8 (30%) clinical diagnosed patients had unilateral involvement, 12 (80%) confirmed diagnosed patients and 19 (70%) clinical diagnosed patients had bilateral involvement. 12 (80%) confirmed diagnosed patients and 15 (65%) clinical diagnosed patients had multiple “ground-glass opacity” lesions in the lung (Table 2 and Figure 2). These clinical manifestations indicated that most COVID-19 infected HD patients are in mild conditions, which is quite different from previous findings on other patients with comorbidities such as diabetes, hypertension, cardiovascular disease or the elderly [7, 8, 16].

**Table 1.** Clinical characteristics of confirmed and clinical diagnosed patients

|  | Confirmed diagnosis | Clinical diagnosed |
| --- | --- | --- |
| No. of patients | 15 | 27 |
| Sex, no. of cases (%) |  |  |
| Male | 10 (67%) | 15 (56%) |
| Female | 5 (33%) | 12 (44%) |
| Age, years |  |  |
| Median (IQR) | 71 (54-76) | 61 (47-68) |
| HD age, months |  |  |
| Median (IQR) | 32 (14-43) | 35 (16-61) |
| Albumin, g/L |  |  |
| Median (IQR) | 39 (36-44) | 40 (35-46) |
| Primary cause of ESRD |  |  |
| Glomerulonephritis, no. of cases (%) | 8 (53%) | 15 (55%) |
| Hypertensive nephropathy, no. of cases (%) | 5 (33%) | 8 (30%) |
| Diabetic nephropathy, no. of cases (%) | 1 (7%) | 4 (15%) |
| Polycystic kidney disease, no. of cases (%) | 1 (7%) | 0 |

**Table 2.**
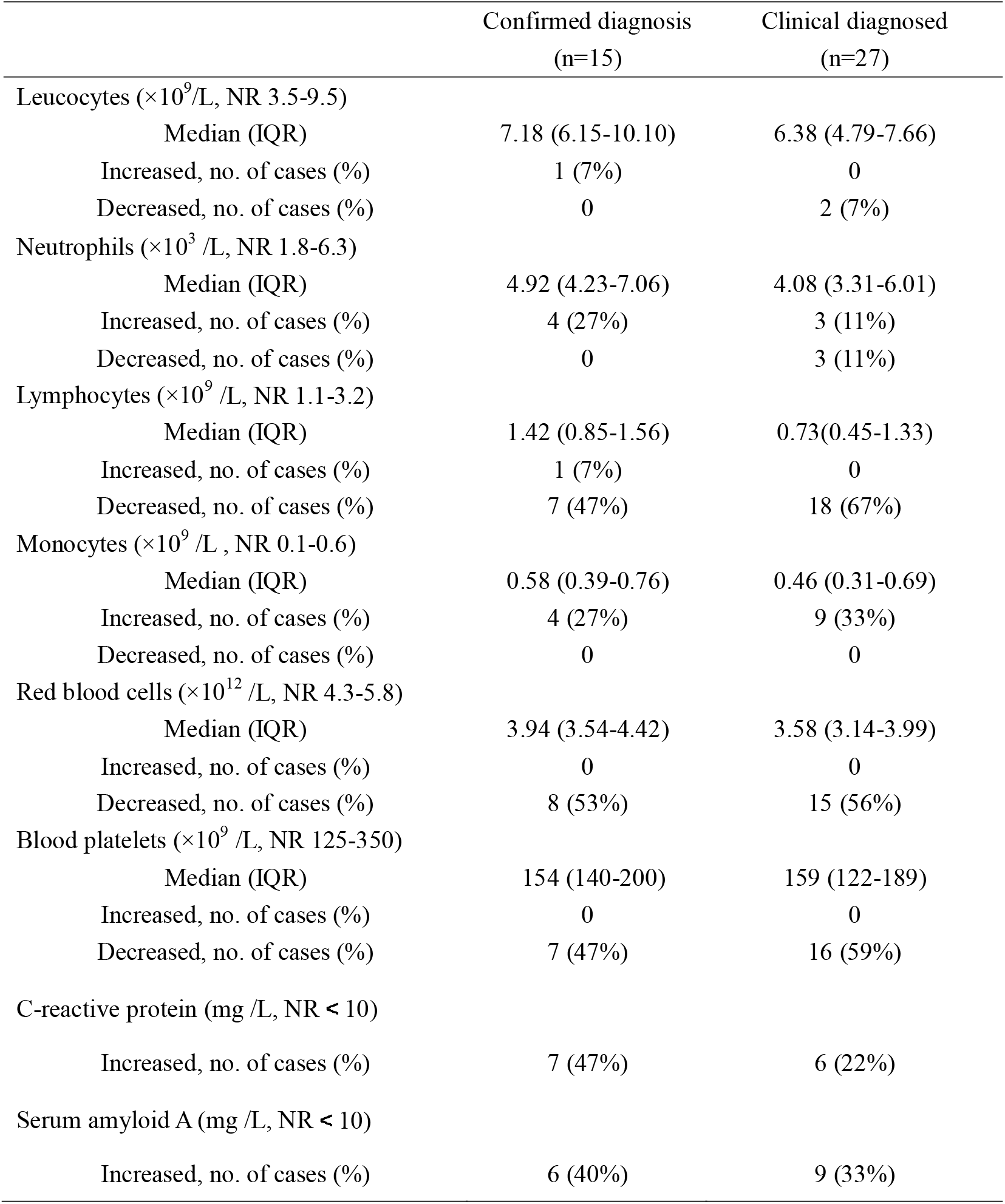
Blood routine findings of confirmed and clinical diagnosed patients

**Table 3.** Clinical symptoms and chest CT results of confirmed and clinical diagnosed patients

|  | Confirmed patients<br>(n=15) | Clinical diagnosed patients<br>(n=27) |
| --- | --- | --- |
| **Symptoms** |  |  |
| Fever, no. of cases (%) | 2 (13%) | 2 (7%) |
| Fatigue, no. of cases (%) | 2 (13%) | 1 (4%) |
| Dry cough, no. of cases (%) | 3 (20%) | 0 |
| Chest pain, no. of cases (%) | 0 | 1 (4%) |
| Nauseating, no. of cases (%) | 1 (7%) | 1 (4%) |
| **Chest CT** |  |  |
| Unilateral pneumonia, no. of cases (%) | 2 (13%) | 8 (30%) |
| Left, no. of cases (%) | 1 (7%) | 2 (7%) |
| Right, no. of cases (%) | 1 (7%) | 6 (22%) |
| Bilateral pneumonia, no. of cases (%) | 12 (80%) | 19 (70%) |
| No pneumonia, no. of cases (%) | 1 (7%) | 0 |
| Multiple “ground-glass opacity” lesions,<br>no. of cases (%) | 12 (80%) | 15 (65%) |

**Table 4.** Epidemiological and clinical characteristics comparisons between HD patients and other populations

|  | HD patients<br>(n=230) | Medical staff<br>(n=33) | General population<br>(n=8837300) | Special group #<br>(n=64) [17] |
| --- | --- | --- | --- | --- |
| No. of infection (%) | 42 (18.3%) | 4 (12.1%) | 49986 (0.6%)[5] | / |
| No. of mild case (%) | 37 (88.1%) | 4(100%) | 43553 (87.1%) | 38 (59.4%) |
| No. of ICU cases (%) | 3 (7.1%) | 0 | 4003(8.0%) | 26(40.6%) |
| No. of deaths (%) | 2 (4.7%) | 0 | 2430(4.9%) | 6 (9.4%) |

#: Indicates COVID-19 infected patients with other comorbidities such as diabetes, hypertension, old age, et al.

**Figure 2.**
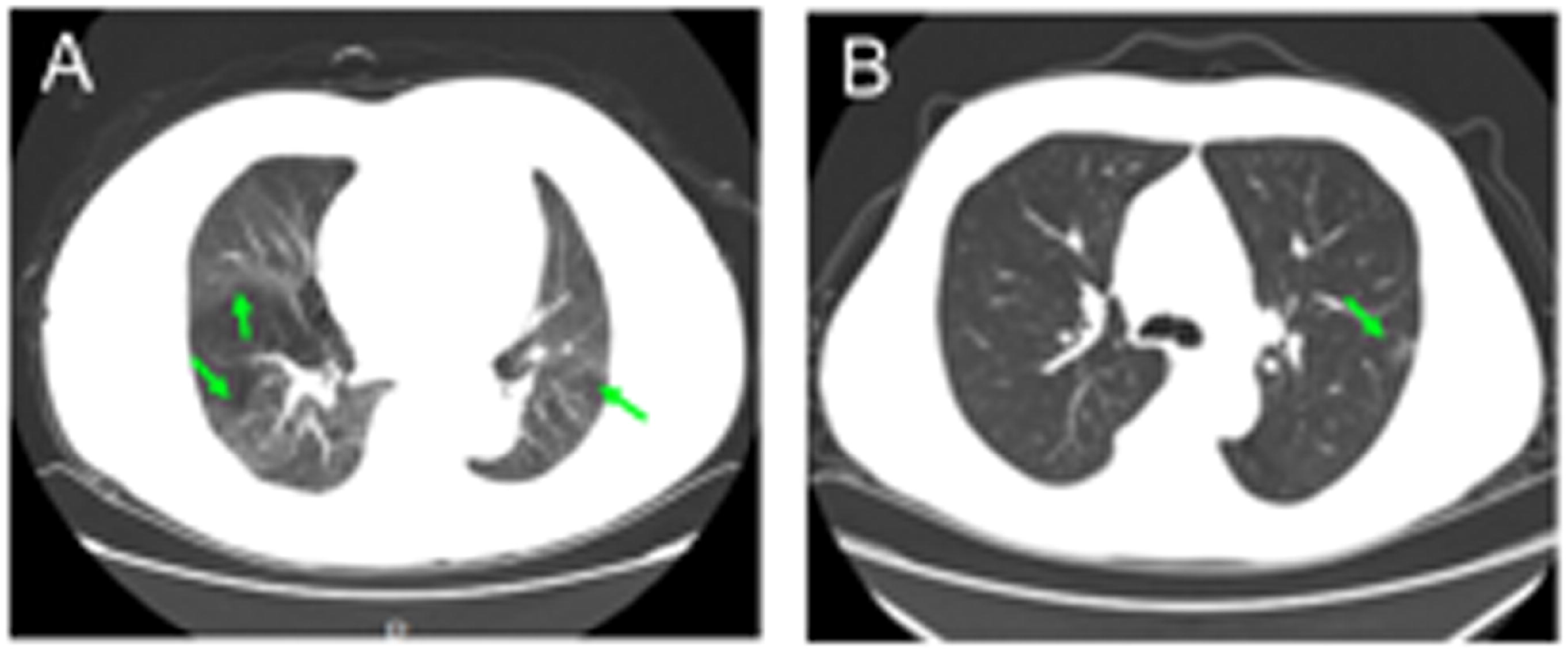
Representative images of chest CT scans (the transverse plane). Patients with COVID-19 had unilateral or bilateral pneumonia. The multiple “ground-glass” lesions were the characteristics of COVID-19 pneumonia. The green arrows represent the sites of lesions.

The immune system plays an essential role in protecting host from pathogens invasion. Once SARS-CoV-2 infected the host’s respiratory tract, it multiplies in cells of the airway, triggering extensive immune activation and releasing massive proinflammatory cytokines. This “cytokine storm” effects are responsible for severe conditions or even eventually lead to death in COVID-19 patients [16]. Above results indicated that patients with SARS-CoV-2 infection were in mild condition. We speculated that this might be related to the compromised immunity in HD patients. To test this hypothesis, we enumerated the absolute numbers of T cells, NK cells, as well as B cells in PBMCs of HD patients in the presence of absence of SARS-CoV-2 infection. We found that the numbers of T cells, Th cells, killer T cells, NK cells and B lymphocytes in PBMCs of HD patients were very low compared with those of non HD patients. Those numbers in HD patients with SARS-CoV-2 infection was further decreased (Figure 3A). Similar to the numbers of lymphocytes, we also observed that the serum levels of IL-4, IL-6, and TNF-α in non HD patients with SARS-CoV-2 infection were significantly higher than the normal level, while the serum levels of these cytokines in HD patients with or without SARS-CoV-2 infection are significantly lower than those in non HD patients with SARS-CoV-2 infection (Figure 3B). These results suggest that HD patients have compromised immune system, which may be detrimental for mounting effective anti-viral responses, but beneficial for limiting tissue damages by dampening the cytokine release.

**Figure 3.**
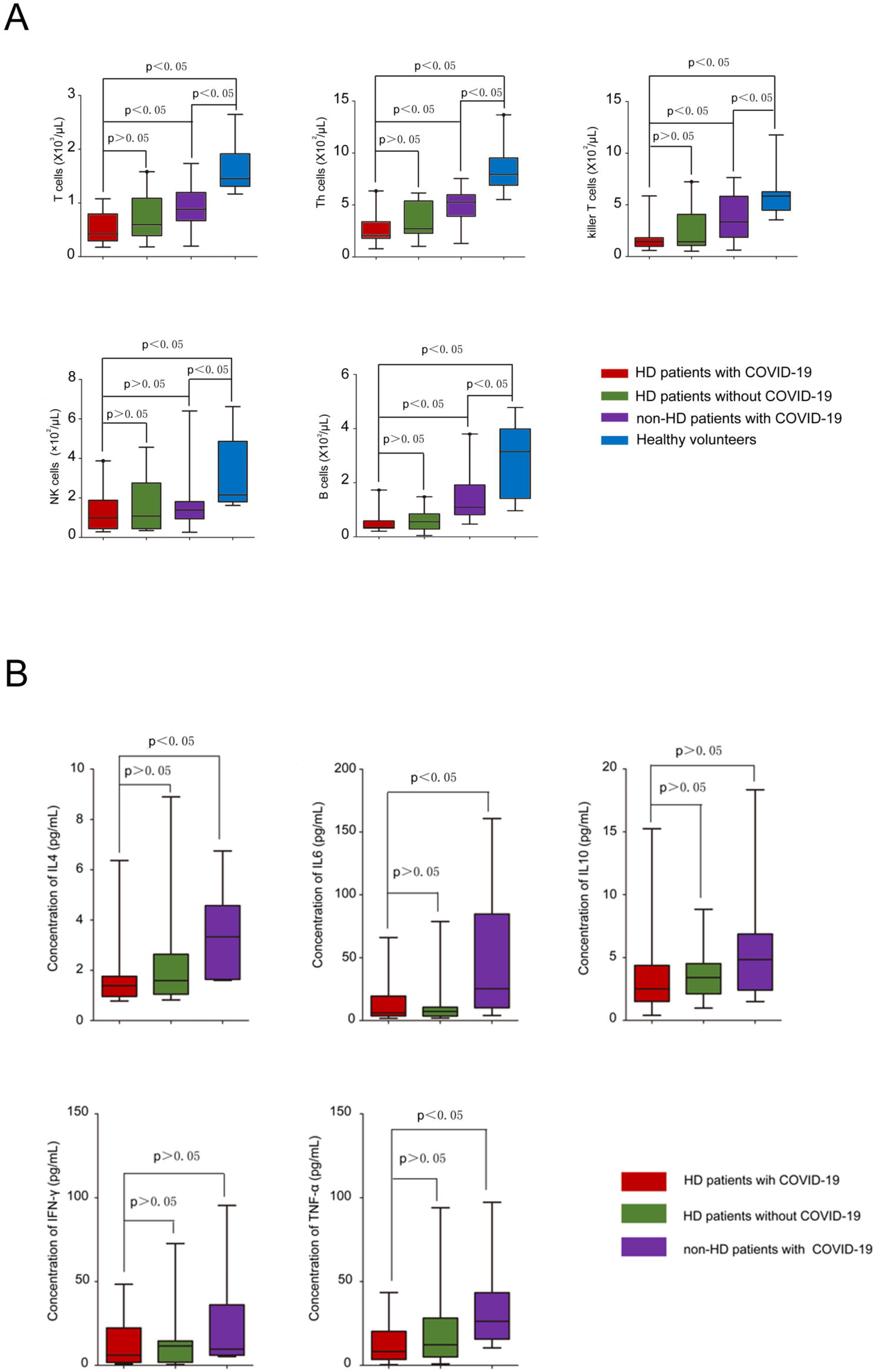
**A**: The frequency of immunocytes in the PBMCs of patients with or without COVID-19. The proportion of T cell, Th cells, killer T cells and B cells of HD patients decreased significantly than other groups. Meanwhile, the proportion of these cells in patients with COVID-19 was decreased than healthy volunteers group. **B:** The serum level of cytokines in indicated patients. The values of IL4, IL6 and TNFα in non-HD COVID-19 patients were significantly higher than those in HD patients with COVID-19.

## Discussion

COVID-19 epidemic, caused by SARS-CoV-2, has swept many countries and regions around the world and sparked an alarming of pandemic by the World Health Organizatino [4]. Previous epidemiological survey showed that the elderly or patients with comorbidities were more susceptible to COVID-19, and the incidence of severe cases and the risk of death were high [16, 18-20]. However, there is no report about the impacts of COVID-19 epidemic on HD patients. HD patients are distinct different cluster from other population because: 1. They make up a large scale cluster; 2. They usually receive concentrated dialysis treatment in a large space; 3. Their immune functions are compromised; 4. Once they are infected, they may become potential “super-spreaders”. Considering these natures, HD patients and HD centers should be given priority in epidemic prevention and control. According to our investigation on single center, the infection rate of HD patients in the COVID-19 epidemic is indeed much higher than that of other population, and the staffs in HD center are also at high risk to infection.

Reviewing the epidemic situation in our center can bring some important experiences and lessons. The COVID-19 epidemic initially emerged in our center on January 14, 2020, but until January 21, actions were taken by our center to stand up to, we must admitted that the best time has missed. Nevertheless, series of measures including upgrading prevention and protection, quarantine and isolation, seem to be effective to contain the epidemic, but the most critical means we think is thoroughly finding out the infected cases by repeated screening, which is mainly based on the results of chest CT scan.

During the outbreak of COVID-19 in our center, 13 episodes of death were recorded, with a mortality of 5.65%, which is far higher than that of the same period in history. The high mortality is obviously related to the epidemic situation. However, it was found that only 2 deaths were pneumonia directly related. The main causes of death were cardiovascular and cerebrovascular complications or hyperkalemia, which was presumed due to the reduction of dialysis times for fear of contracting the virus. It should be noted that although HD patients are highly susceptible to COVID-19, the infection for them is likely less severe or fatal. Actually, only 3 of 42 infected patients were admitted to ICU. Some infected patients even have no obvious clinical symptoms. These suggest that in the face of the COVID-19 epidemic emerging, measures of prevention and protection certainly should be taken to avoid infection, but the sufficient dialysis remains the essential for patients to survive the epidemic. During the outbreak of SARS in 2003, it was observed that the severity and mortality of HD patients infected with SARS-CoV were similar to other infected population, but the duration of shedding virus through stool or breath was significantly longer than other patients [21]. Considering the high biological similarity between SARS-CoV-2 and SARS-CoV, we estimate that HD patients infected with SARS-CoV-2 will take longer to clear the virus, and these patients may need longer quarantine period to prevent the spread of infection.

Previous studies have shown that SARS-CoV-2 infection can reduce the number of lymphocytes in patients, but the level of inflammatory cytokines in vivo increases significantly. Cytokine storm may be the key cause of the worsened condition and even death [16]. We tested the frequency of immune cells in the PBMCs of patients with and without SARS-CoV-2 infection and the level of cytokines in the body of patients. The results showed that compared with the general population, the T cells, Th cells, killer T cells, as well as NK cells was reduced remarkably in PBMCs of HD patients, irrespective of SARS-CoV-2 infection. In addition, the serum level of serial cytokines of IL-4, IL-6, TNF-α in SARS-CoV-2 infected HD patients remain relatively lower in comparison with non-HD patients with SARS-CoV-2 infection. This indicates that the impaired immune system seems to be unable to mount effective cellular immune response upon the invasion of SARS-CoV-2, thus results in no cytokine storm and no severe organs damage. In the current clinical guidelines and practice for COVID-19 therapy, glucocorticoids are recommended and usually prescribed [7, 12]. However, with regard to the management of COVID-19 in HD patients, we suggest the administration of glucocorticoids should be prudent since the immune system in HD patients have already been suppressed [22-24].

## Data Availability

All data generated or analyzed in this study are included in this published article.

## Acknowledgments

This work was supported by the grants from the National Natural Science Foundation of China (#81370800 to Dr. Huiming Wang, # 81800615 to Dr. Yiqiong Ma), the Key Project on Science and Technology Innovation of Hubei Province (#2019ACA137 to Dr. Huiming Wang), the Key Project on Health Commission of Hubei Province (#WJ2019Z011 to Ming Shi).

